# Hypervirulent extensively-drug resistant (XDR) *Klebsiella pneumoniae* associated with complicated urinary tract infection in Northern India

**DOI:** 10.1101/2021.05.15.21256863

**Authors:** Parinitha Kaza, Xavier Basil Britto, Jaspreet Mahindroo, Stephen Baker, To Nguyen Thi Nguyen, Ravimohan Suryanarayana Mavuduru, Balvinder Mohan, Neelam Taneja

## Abstract

*Klebsiella pneumoniae* associated with hospital acquired infections in South Asia are frequently extensively-drug resistant (XDR), making treatment and control problematic. It is important to understand the epidemiology and genetic structure of XDR *K. pneumoniae* and to determine their potential to be hypervirulent (hv) through the presence of siderophores. Here, we characterized the genomes of 20 colistin-resistant XDR *K. pneumoniae* isolated from 16 patients with complicated UTI over a six-month period in a healthcare facility in northern India. The 16 organisms comprised five STs: ST14 (10/20), ST147 (5/20), ST231 (3/20), ST2096 (1/20), and ST25 (1/20). Notably, several patients admitted to a single ward were infected with the same ST, potentially indicating a common infection source. Additionally, some patients had recurrent infections with multiple STs that were circulating concurrently in a particular ward, providing further evidence for hospital transmission. Beta lactamase genes (*bla*_CTX-M-1_, *bla*_SHV_, *bla*_LEN_, and *bla*_AMP-H_) were present in all isolates and the *bla*_NDM,_ *bla*_OXA1-90_, and *bla*_OXA48_ carbapenemases were present in 17, 18, and 3 isolates, respectively. Disruption of *mgrB* with various IS elements was identified in six organisms and was the most common mechanism of colistin resistance. The most frequent K type was K2 (11/20), followed by K10, K51, and K64. Notably, we identified one XDR convergent hypervirulent *K. pneumoniae* (hvKp) associated with prolonged hospitalisation (*iuc+ybt+ESBL+*OXA-1, OXA-48), belonging to ST2096. Our data suggest that convergent XDR-hvKp is circulating in our healthcare facility. We speculate that such organisms may have outbreak potential, warranting more effective antimicrobial stewardship and better infection control strategies.

## Introduction

There is growing concern regarding the disease burden associated with the ESKAPE group of pathogens (*Enterococcus faecium, Staphylococcus aureus, Klebsiella pneumoniae, Acinetobacter baumannii, Pseudomonas aeruginosa*, and *Enterobacter* species). These bacteria are frequently multi-drug resistant (MDR) and are a leading cause of hospital-acquired infections internationally (1). More specifically, *K. pneumoniae* is a major emerging pathogen and has been implicated in a wide variety of infections, including meningitis, pneumonia, sepsis, intra-abdominal infections, skin and soft tissue infections, and urinary tract infections (UTIs) (2). *K. pneumoniae* that are acquired in hospitals in South Asia are frequently extensively-drug resistant (XDR) or pan-drug resistant (PDR), making them highly problematic (3). An increase in XDR and PDR *K. pneumoniae* has become a global concern, and the World Health Organization (WHO) has placed carbapenem and third-generation cephalosporin-resistant *K. pneumoniae* on the priority list of pathogens requiring new antimicrobials (4).

The most dangerous form of *K. pneumoniae* are associated with invasive disease and were first isolated from liver abscesses; these invasive organisms are considered hypervirulent (hvKp) (5). HvKp variants are more commonly community-acquired, but their capacity to infect healthy individuals may also facilitate their transmission in healthcare facilities. Consequently, hvKp infections are becoming more prevalent and associated with increasing mortality (6). The molecular basis of hvKp is not implicit, but isolates that carry the salmochelin, enterobactin, and aerobactin siderophores appear to be better adapted for systemic spread in humans. Notably, the *iuc* locus (*iucABCD*), which encodes aerobactin, is located on a plasmid and can theoretically be horizontally transferred from one organism to another. Thankfully, hvKp isolates are generally susceptible to the majority of clinically relevant antimicrobials, but there are emerging reports of hvKp isolates with MDR phenotypes (7). The combination of extreme AMR phenotypes (MDR, XDR, and carbapenem resistance) and hypervirulence in *K. pneumoniae* represent a major threat to public health.

*K. pneumoniae* is the second most common cause of UTI in our tertiary care centre in Northern India, accounting for 25% of all UTIs; 13.9% of these are XDR (8). The antimicrobial susceptibility of these organisms determines that colistin is now the last resort drug for the treatment of XDR *K. pneumoniae* infections (9). However, the reported prevalence of colistin non-susceptibility in MDR *K. pneumoniae* of 18.5% in our centre is also high (8). The presence of the *mcr* gene and variants, mutations in the *PhoPQ, PmrAB* two-component systems, *mgrB* gene mutations in these organisms, lead to colistin resistance and compromise the potential efficacy of this vital drug (10-12).

With the potential health implications of complicated UTIs caused by MDR and XDR organisms, it is essential to better understand their epidemiology and genetic composition. Whole-genome sequencing (WGS) can help us identify both known and novel mechanisms of antimicrobial resistance, determine sequence types (STs), and aid epidemiological investigations through phylogenetics (12). Here, we characterized the genomes of 20 colistin-resistant XDR *K. pneumoniae* that were isolated from 16 patients with complicated UTI over a six-month period in a tertiary hospital in Northern India.

## Methods

### Study design

We conducted a retrospective study to characterize the genome sequences of 20 colistin non-susceptible uropathogenic XDR *K. pneumoniae* isolated from urine samples of 16 in-patients attending the Postgraduate Institute for Medical education and Research (PGIMER) in Chandigarh. The clinical details of the patients were recorded from the case files. This study was approved by the internal ethics review board of PGIMER.

### Microbiological methods

Urine samples from patients suffering from UTI (Fever >100□ F, burning micturition, and increased frequency of urination) admitted to PGIMER between July 2017 and January 2018 were processed according to the standard operating procedures of the Enteric Laboratory (8). Samples were inoculated on cysteine lactose electrolyte deficient (CLED) media; yellow colonies (lactose fermenting) indicative of *K. pneumoniae* were subjected to Matrix-assisted laser desorption/ ionization-time of flight mass spectrometry (MALDI-TOF) (Bruker Dalton, Germany). Those confirmed as *K. pneumoniae* were subjected to antimicrobial susceptibility testing (AST) and their clinical details were accessed from case records (8).

AST was performed using the Kirby-Bauer disc diffusion method for aminoglycosides [amikacin (30 µg), gentamicin (10 µg)], beta-lactams [piperacillin-tazobactum (100/10 µg)], carbapenems [ertapenem (10 µg), meropenem (10 µg), imipenem (10 µg)], cephalosporins [cefotaxime (30 µg), cefepime (30 µg), cefoparazone-sulbactum (75/30 µg)], co-trimoxazole (1.25/23.75 µg), quinolones [ciprofloxacin (5 µg), norfloxacin (10 µg), nalidixic acid (30 µg)], tetracyclines [tetracycline (30 µg), minocycline (30 µg)], nitrofurantoin (300 µg), tigecycline (15 µg) and fosfomycin (200 µg) (Becton, Dickinson and Company., Sparks, MD, USA) following the recommendations of the Clinical and Laboratory Standards Institute (CLSI) (13). MDR was defined as non-susceptibility to at least one agent in three or more antimicrobial categories and XDR was defined as non-susceptibility to at least one agent in all but two or fewer antimicrobial categories (i.e., bacterial isolates remaining susceptible to only one or two categories). XDR isolates were further tested using the CLSI broth microdilution method to determine phenotypic resistance to colistin (Sigma Aldrich, USA) (14). Isolates with minimum inhibitory concentration (MIC) >=2 µg/mL against colistin were deemed as colistin non-susceptible.

### Whole-genome sequencing

Genomic DNA was extracted from all isolates using the Wizard genomic DNA purification kit (Promega, Madison WI, USA). Briefly, genomic DNA was purified from each isolate and quantified using a Qubit dsDNA high sensitivity assay (ThermoFisher, Waltham, USA). Libraries were prepared using the Nextera XT kit and sequenced using an MiSeq platform (2 x 250 bp, V2 500 cycles) (Illumina Inc., San Diego, CA,USA). The raw sequences were subjected to a quality check and analysis using an in-house pipeline, BacPipe (15). Briefly, reads were quality assessed, trimmed, and *de novo* assembled (SPAdes v 3.10.0) (16). The assemble sequences were subjected to MLST typing, annotation (Prokka v 1.12) (17), and antimicrobial resistance gene prediction using ARG-annot v3 (18). In order to determine the incompatibility type of the plasmids present in the sequences, the data were processed using Plasmid Finder 2.1 (19) and insertion element (IS) families were detected using IS Finder (20). Virulence genes were identified using VFDB (21) and capsular typing was performed using Kleborate v0·3·0 (http://github.com/katholt/Kleborate).

### Phylogenetic Analysis

For phylogenetic comparison, all sequenced isolates along with a reference sequence (21 genome sequences) were passed through the RedDog pipeline V1beta.10.3 (https://github.com/katholt/RedDog). Phylogenetic relationships were inferred using the Maximum Likelihood method based on a General Time Reversible model. A phylogenetic tree was constructed to scale, with branch lengths measured in the number of substitutions per site. A maximum likelihood tree for ST14 (the most common ST in this study) isolates was constructed along with additional genomes available in GenBank downloaded using SRA Toolkit (https://www.ncbi.nlm.nih.gov/sra/). The phylogenetic trees were viewed on iTOL (22).

## Results

### Baseline characteristics

Over the six-month sampling period 775 (3.1%) *K. pneumoniae* were isolated from 25,046 urine samples from patients with a clinically suspected UTI. Following antimicrobial susceptibility testing, 108 (13.9%) MDR *K. pneumoniae* isolates were identified. Twenty of these *K. pneumoniae*, isolated from 16 patients, were non-susceptible to colistin, and latterly determined to be XDR. The majority of these *K. pneumoniae* were isolated from patients admitted to the urology wards (n=12), all of which had structural abnormalities of the urinary tract or had recently undergone urinary tract surgery (Supplementary File 1).

### Antimicrobial resistance gene content and phylogenetic analysis

These 20 colistin resistant *K. pneumoniae* isolates were subjected to WGS. We initially observed the presence of a wide variety of AMR genes (41 individual genes) within these isolates. Specifically, the beta-lactamase genes *bla*_CTXM-1_, *bla*_SHV-1_, and *bla*_AmpH_ were present in all isolates. Furthermore, 17/20 sequences contained *bla*_NDM_, of which 15/17 sequences also contained *bla*_OXA-1_, the remaining 2/17 contained *bla*_OXA-48_. None of the isolates were KPC producers (Fig. 1). With respect to colistin resistance, the WGS data showed absence of *mcr* 1-10 genes and mutations in the *PhoPQ and PmrAB* two-component systems. However, 8/20 isolates contained deletions/mutations in the *mgrB* genes, which included a disruption with IS110 (most common)/ISKpn26 and Cys28Gly (Table 1).

**Table 1.**
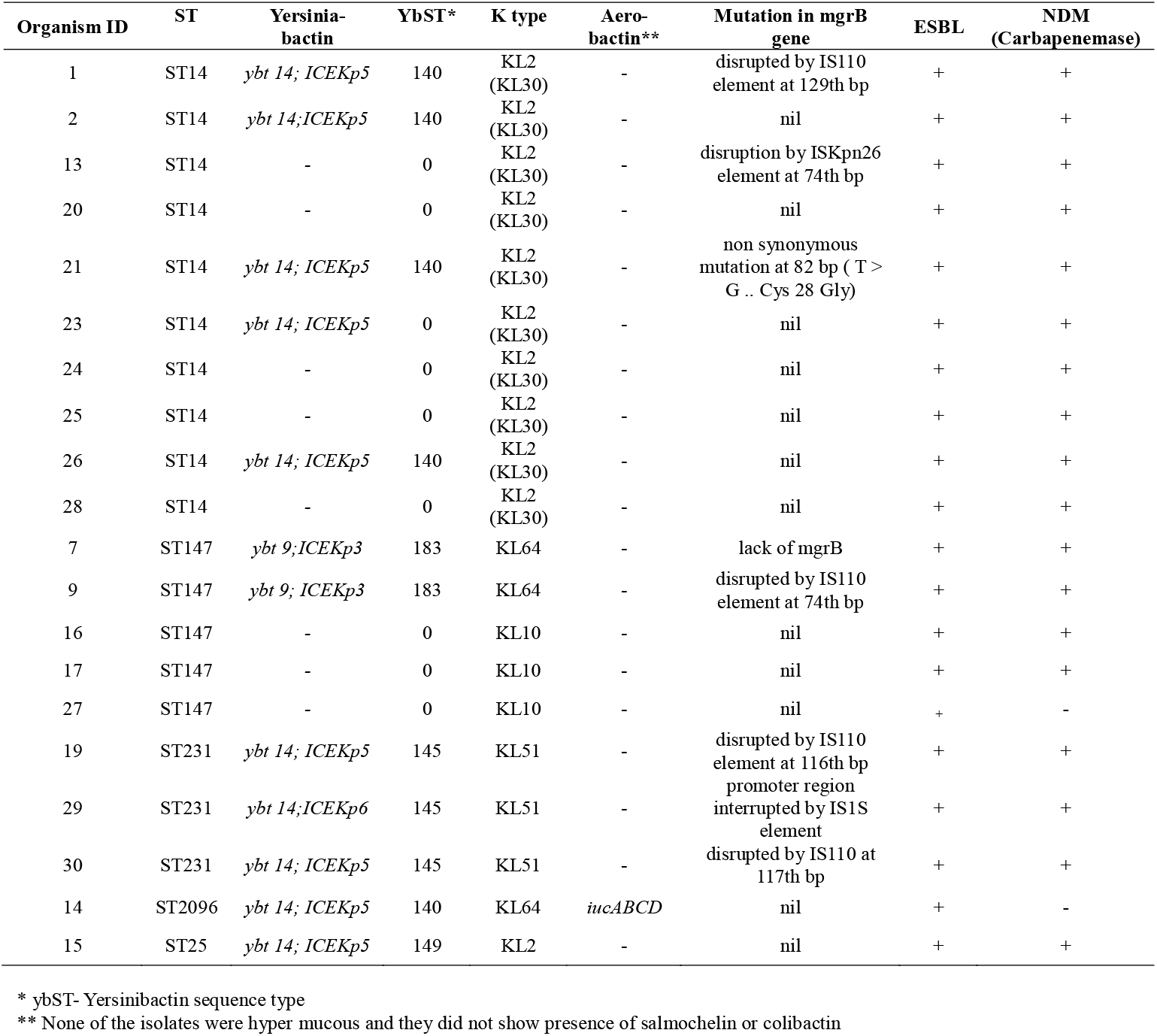
The distribution of AMR and virulence determinants in sequenced organisms

**Figure 1.**
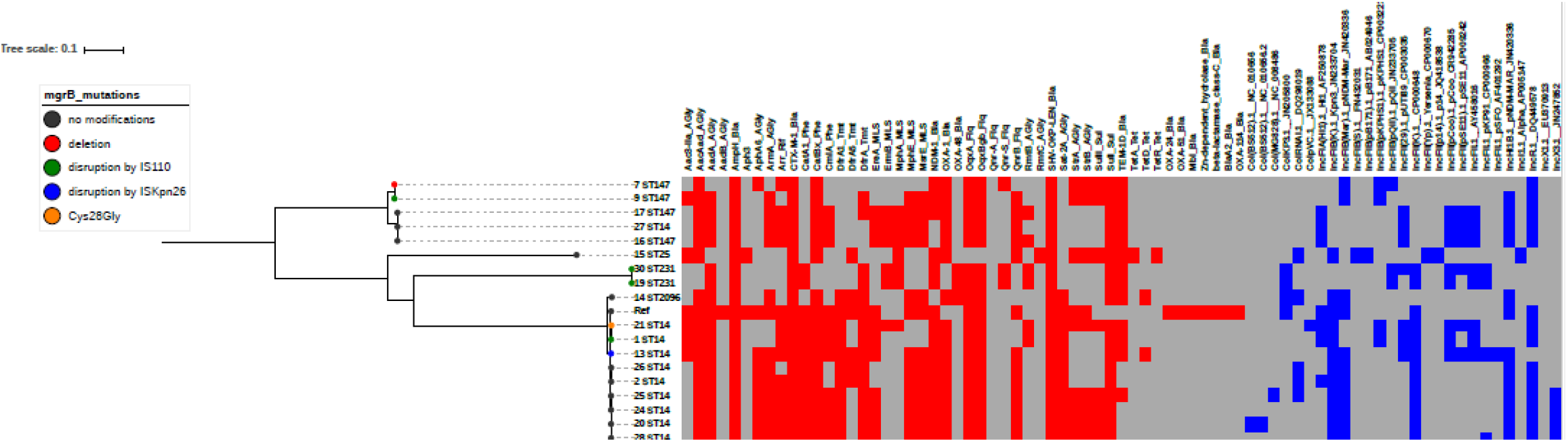
Phylogenetic structure of colistin resistant *K. pneumoniae* isolates from this study. A phylogenetic tree of *K. pneumoniae* isolates from this study was constructed based on core SNP alignment using RedDog pipeline and viewed on iTOL with a heat-map showing AMR genes (red) and plasmids replicons (blue).

We identified five different STs; the most common was ST14 (n=10), followed by ST147 (n=5), ST231 (n=3), ST2096 (n=1), and ST25 (n=1) (Fig. 1). We observed two phylogenetically distinct variants of ST147, of which three isolates exhibited an identical compendium of AMR genes and an indistinguishable plasmid profile. Furthermore, two ST231 isolates additionally shared an identical plasmid with the same AMR genes. Multiple STs were isolated from two patients (P3 and P15).Patient P3, with a bulbar urethral stricture, had recurrent UTIs with ST147 and ST25 and patient P15, with autosomal dominant polycystic kidney disease (ADPKD) and a right hydro-ureter, had multiple UTI episodes caused by ST14, ST147, ST231 and ST25. We surmised that ST147 and ST231 were likely circulating concurrently in the urology ward (Supplementary File 1, Fig. 2).

**Figure 2.**
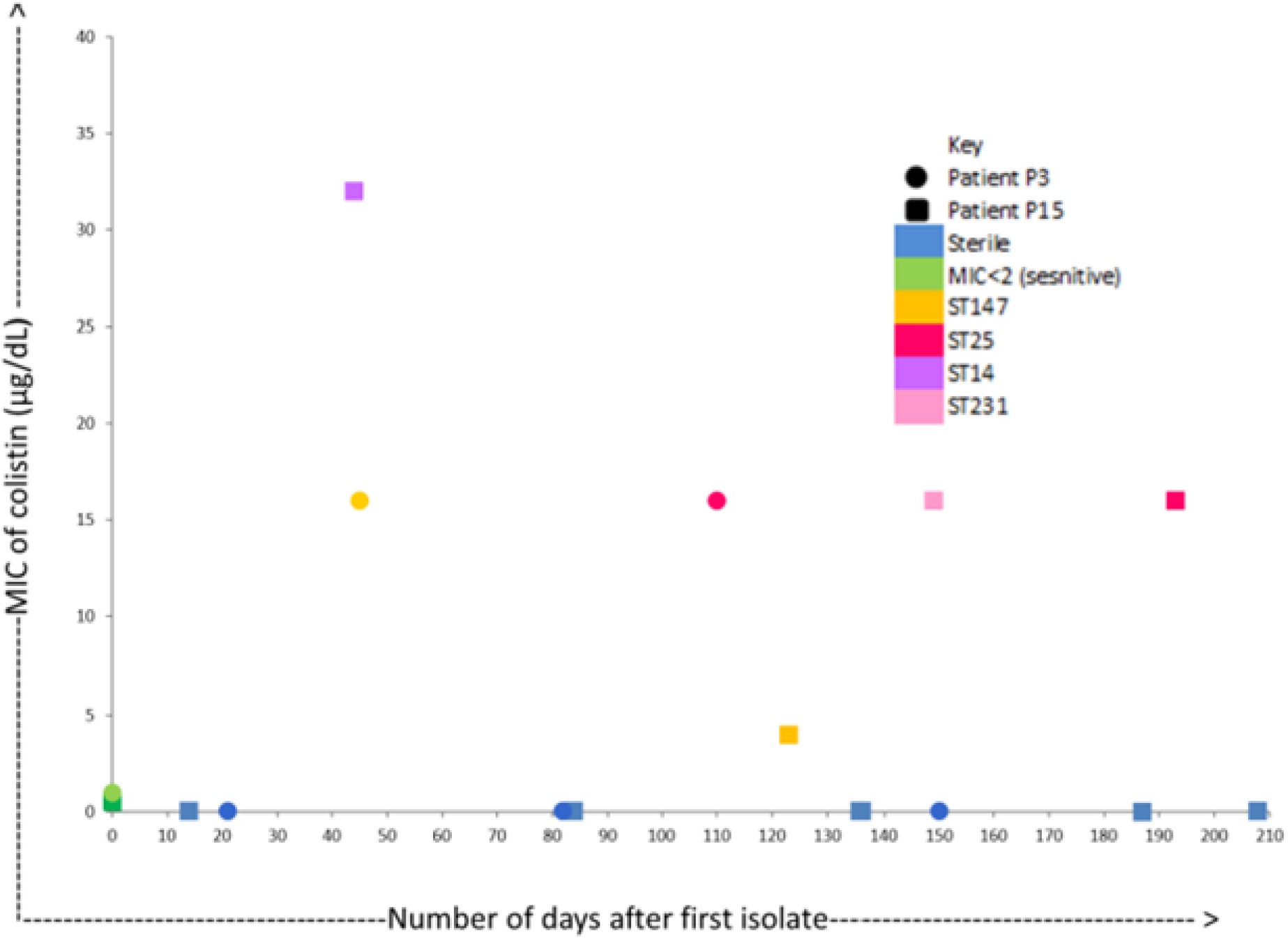
Patients with recurrent UTI due to colistin resistant *K. pneumoniae*. A scatter plot of multiple *K. pneumoniae* STs isolated from two patients showing their colistin MIC on x-axis and no of days on y-axis.

As ST14 was the most common genotype we constructed an independent phylogeny combining our ST14 sequences with sequences of ST14 *K. pneumoniae* from public databases (Fig. 3). The phylogenetic tree from core SNPs formed three independent clades; the isolates originating from India clustered into a single clade. We again detected a wide array (n=35) of AMR genes in these ST14 isolates, which were associated with resistance to several antimicrobial classes including aminoglycosides, trimethoprim, fluoroquinolones, phenicols, sulphonamides, streptomycin, tetracycline, fosfomycin, rifampin, 16S *rRNA* methylases, and beta lactams. Genes encoding resistance to aminoglycosides were present in all Indian isolates, as was the *fosA* gene coding resistance to fosfomycin. Additionally, the *bla*_NDM*1*_, *bla*_OXA1_ and *bla*_CTXM_ carbapenemases were present in all isolates. Other detected ESBL genes included *bla*_TEM_ in two isolates and *bla*_SHV-1_ in six isolates. Notably, the ST14 *K. pneumoniae* isolates from outside of India (e.g., USA) had only a limited number of AMR genes in comparison to those originating from India (Supplementary File 2, Fig. 3).

**Figure 3.**
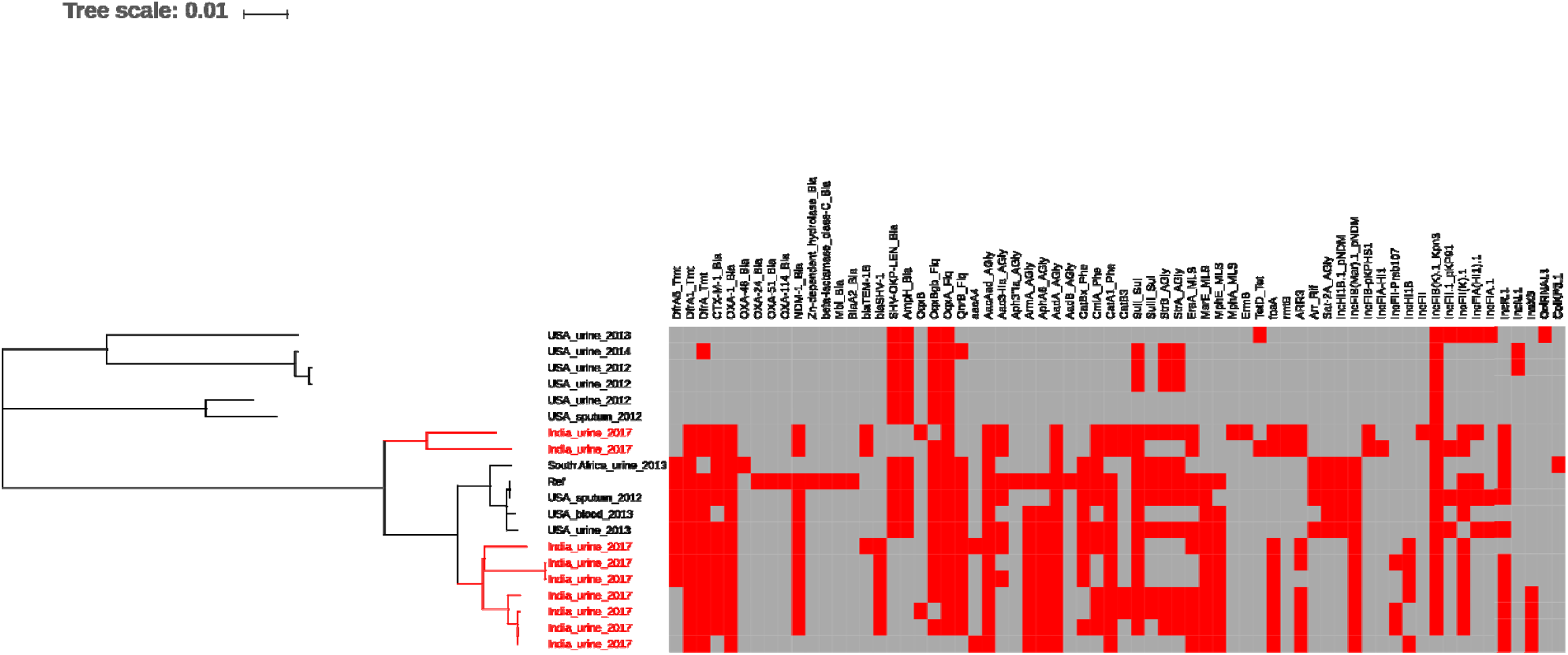
Phylogenetic structure of ST14 *K. pneumoniae* isolates. A core SNP phylogenetic tree of ST14 K. pneumoniae with published genomes was constructed using RedDog pipeline and viewed on iTOL with a heat map showing AMR genes (red)

### Virulence gene analysis and capsular typing

We lastly assessed the composition of virulence-associated genes. The *ybt* locus was the most common being present in 50% (10/20) of isolates and found in multiple STs [ST231 (n=3), ST2096 (n=1), ST14 (n=5), and ST147 (n=2)]. A single hypervirulent *K. pneumoniae* possessing the *iuc* locus was found and belonged to ST2096, a single locus variant of ST14. The plasmid in this isolate also carried an ESBL gene and carbapenemases (*bl*a_OXA-1_ and *bla*_OXA-48_). Consequently, we determined this ST2096 isolate to be a convergent XDR-hvKp variant. The majority (7/10) of isolates possessing *ybt* were also found to be colistin resistant, with corresponding mutations in *mgrB*. All isolates with *ybt* locus also possessed ESBL genes and 18/20 were metallo-β lactamase producers. Additionally, all isolates contained the *fepG* (iron enterobactin) and *entS* (enterobactin) siderophores, which also contribute to virulence. No hypermucous variants, or isolates containing salmochelin or colibactin were identified (Table 1 and Supplementary File 1). The most common K type among these isolates was K2 (11/20), followed by K10, K51, and K64.

## Discussion

*K. pneumoniae* is the second most common organism associated with complicated with UTIs after *E. coli*, accounting for ∼20% of cases at our centre (unpublished data). Crucially, the prevalence of MDR in *K. pneumoniae* is six times greater than observed in *E. coli* and UTIs caused by MDR *K. pneumoniae* are associated with increased morbidity (8). Colistin resistance is also emerging in India (23, 24), and we also identified organisms that were non-susceptible to colistin. Colistin resistance in *K. pneumoniae* is commonly associated with modification of lipid A, mutations in efflux pumps and the plasmid associated *mcr* genes (25). We did not detect any *mcr* genes or evidence for mutation in *Pmr*A*/Pmr*B, *Pho*P*/Pho*Q *ram*A or *ccr*B. We detected mutations in *mgrB*, which encodes a small transmembrane protein that mediates negative feedback on the *PhoQ/PhoP* system (26, 27). Mutations in *mgr*B explain 40% of colistin resistance observed here, but other mechanisms like accumulation of capsular polysaccharide or efflux pumps may also contribute to colistin resistance in *K. pneumoniae* (28).

ST14 was the predominant ST in this study. This is in accordance to the study by Long *et al*., who also found ST14 to be common in India (29). A further study from Vellore detected ST231, ST14 and ST147, which is again comparable to the distribution of STs observed here. By combining data from comparative analysis of ST14 isolates from aforementioned study in Vellore (28), the USA (29), and Pakistan (30), and our data, we observed that Indian isolates possess more AMR genes than the organisms form other locations. Additionally, the ST14 isolates from India clustered into a single clade, indicating a common origin, which is concerning given that they have the capacity to cause UTIs and bloodstream infections.

The majority of isolates here possessed enterobactin (*entS*), and half had siderophores such as yersinibactin (*ybt*), both of which contribute to the virulence (31). The iron scavenging ability of enterobactin can be limited by the binding of host lipocalin-2 to aferric enterobactin. The presence of *ybt* aids in overcoming this defence mechanism and facilitates bacterial invasion (32, 33). Here, all isolates possessing *ybt* were either ESBL or carbapenemases producers. The *ybt* locus is mobilised by a self-transmissible integrative conjugative element (ICE), which can also carry additional virulence determinants (*iro, clb*) (32). We identified a single ST2096 *K. pneumoniae* with aerobactin *(iucABCD*), *ybt* and ESBL, OXA-1 and OXA48 that, that we defined as a convergent XDR-hypervirulent variant. This organism was isolated from a patient with recurrent UTI and renal failure associated with urinary tract abnormalities. The patient was treated with aminoglycosides, cephalosporins, cotrimoxazole, and colistin during his 23 days of hospitalization. Despite antimicrobial therapy, recovery was prolonged and bacterial cultures continued to remain positive 10 days after corrective surgery. Concerningly, the presence of convergent hvKp have been reported from various parts of India (24) and Wyres *et al*. described Indian hvKp belonging to ST2096 and ST231 (7). Generally, hvKp is mostly confined to community transmission, but we speculate that the virulence plasmid can integrate into classical hospital XDR *K. pneumoniae* and trigger major outbreaks.

The main concern with highly resistant and hvKp inducing UTIs is that they are associated with high morbidity. Many patients exhibit clinical evidence of urosepsis, even in infections caused by non-hvKp. Here, we observed that several patients admitted to a single ward were infected with the same ST, indicating that these infections may have been acquired from a common source. Concurrently, some patients had recurrent infections with multiple STs that were circulating on a specific ward at that time, providing further evidence for hospital transmission. Patients with recurrent UTIs often have prolonged hospital stays and have generally been exposed to multiple antimicrobials, including carbapenems and colistin (8). MDR *K. pneumoniae* that circulate in hospitals are a major concern as resistance genes are principally plasmid-mediated, and their ability to acquire further resistance genes gives them potential to become XDR and PDR. This scenario could lead to the development of more serious outbreaks associated with higher morbidity and mortality.

With the majority of our colistin-resistant isolates being ESBL and NDM-1 producers, there are limited options to treat these XDR infections (7). Therefore, we need new approaches to prevent and infections caused by these organisms. Formulating appropriate antimicrobial and infection control policies, compliance with hand hygiene, barrier nursing, and antimicrobial stewardship may aid in slowing the emergence of these highly resistant pathogens. Periodic surveillance of hospital wards to assess the circulating organisms and their antimicrobial susceptibility profiles should better support the design of empirical therapies.

## Conclusions

This study provides new insight into the genes facilitating of AMR and virulence in *K. pneumoniae* associated with UTIs in India. Our data suggest that convergent MDR-hvKp is present in our healthcare facility; we hypothesise that such organisms have outbreak potential, warranting more effective antimicrobial stewardship and better infection control strategies in India.

## Data Availability

Raw Illumina reads of Klebsiella pneumoniae isolates in this study have been deposited in the Sequence read archive (SRA), NCBI under the bio project PRJNA530687 (https://www.ncbi.nlm.nih.gov/bioproject/PRJNA530687). Additionally, 13 published genomes of K. pneumoniae from study accession numbers PRJNA376412 and PRJNA287968 were also included in the analysis. All the accession numbers, project numbers, and their corresponding metadata are listed in Table S1.

## Data Availability

Raw Illumina reads of *Klebsiella pneumoniae* isolates in this study have been deposited in the Sequence read archive (SRA), NCBI under the bio project PRJNA530687 (https://www.ncbi.nlm.nih.gov/bioproject/PRJNA530687). Additionally, 13 published genomes of *K. pneumoniae* from study accession number PRJNA376412 (29) and PRJNA287968 (34) were also included in the analysis. All the accession numbers, project numbers and their corresponding metadata are listed in Table S1.

## Competing interests

None to declare

## Author contributions

PK Sample collection, processing, data analyses, manuscript writing

XBB Whole genome sequencing, MLST

JM Phylogenetic and MLST analyses, manuscript writing

SB Manuscript writing and editing

TN Capsular typing of *K. pneumoniae*, virulence determinants analyses

RSM clinical details

BM Editing manuscript

NT Conceptualization, Supervision, manuscript writing and editing

## Funding

Stephen Baker is funded by Wellcome senior research fellowship (215515/Z/19/Z).

## Ethical considerations

This work was a part of the MD Thesis of Dr. Parinitha Kaza, the study was conducted at PGIMER Chandigarh, after the approval of the Institute Ethics Committee of PGIMER reference no NK/3761/MD/386; vide letter no INT/IEC/2017/1009 dated 12.10.2017. The study was also approved from Institute Collaborative Research Committee of PGIMER vide letter no 79/227-Edu-18/4997 dated 12.12.2018.

## Notes

### Competing Interest Statement

The authors have declared no competing interest.

## References

1. Santajit S, Indrawattana N. Mechanisms of Antimicrobial Resistance in ESKAPE Pathogens. BioMed research international. 2016;2016:2475067.

2. Paczosa MK, Mecsas J. Klebsiella pneumoniae: Going on the Offense with a Strong Defense. Microbiol Mol Biol Rev. 2016;80:629–61.

3. Magiorakos A-P, Srinivasan A, Carey RB, Carmeli Y, Falagas ME, Giske CG, et al. Multidrug-resistant, extensively drug-resistant and pandrug-resistant bacteria: an international expert proposal for interim standard definitions for acquired resistance. Clinical microbiology and infection : the official publication of the European Society of Clinical Microbiology and Infectious Diseases. 2012;18(3):268–81.

4. WHO publishes list of bacteria for which new antibiotics are urgently needed [press release]. GENEVA 2017.

5. Liu YC, Cheng DL, Lin CL. Klebsiella pneumoniae liver abscess associated with septic endophthalmitis. Archives of internal medicine. 1986;146(10):1913–6.

6. Russo T, Marr C. Hypervirulent Klebsiella pneumoniae. Clin Microbiol Rev. 2019;32(e00001–19).

7. Wyres KL, Nguyen TN, Lam MM, Judd LM, van Vinh Chau N, Dance DA, et al. Genomic surveillance for hypervirulence and multi-drug resistance in invasive Klebsiella pneumoniae from south and southeast Asia. bioRxiv. 2019:557785.

8. Kaza P, Mahindroo J, Veeraraghavan B, Mavuduru RS, Mohan B, Taneja N. Evaluation of risk factors for colistin resistance among uropathogenic isolates of Escherichia coli and Klebsiella pneumoniae: a case-control study. J Med Microbiol. 2019;68(6):837–47.

9. Lee CR, Lee JH, Park KS, Kim YB, Jeong BC, Lee SH. Global Dissemination of Carbapenemase-Producing Klebsiella pneumoniae: Epidemiology, Genetic Context, Treatment Options, and Detection Methods. Frontiers in microbiology. 2016;7:895.

10. Liu Y-Y, Wang Y, Walsh TR, Yi L-X, Zhang R, Spencer J, et al. Emergence of plasmid-mediated colistin resistance mechanism MCR-1 in animals and human beings in China: a microbiological and molecular biological study. The Lancet Infectious Diseases. 2016;16(2):161–8.

11. Haeili M, Javani A, Moradi J, Jafari Z, Feizabadi MM, Babaei E. MgrB Alterations Mediate Colistin Resistance in Klebsiella pneumoniae Isolates from Iran. Front Microbiol. 2017;8(2470).

12. Pecora ND, Li N, Allard M, Li C, Albano E, Delaney M, et al. Genomically Informed Surveillance for Carbapenem-Resistant Enterobacteriaceae in a Health Care System. mBio. 2015;6(4):e01030.

13. CLSI. Performance Standards for Antimicrobial Susceptibility Testing. Clinical and Laboratory Standards Institute antimicrobial susceptibility testing standards. 2017(27th Edition).

14. Sun K, Xu X, Yan J, Zhang L. Evaluation of Six Phenotypic Methods for the Detection of Carbapenemases in Gram-Negative Bacteria With Characterized Resistance Mechanisms. Ann Lab Med. 2017;37(4):305–12.

15. Xavier BB, Mysara M, Bolzan M, Ribeiro-Gonçalves B, Alako BTF, Harrison P, et al. BacPipe: A Rapid, User-Friendly Whole-Genome Sequencing Pipeline for Clinical Diagnostic Bacteriology. iScience. 2020;23(1):100769.

16. Bankevich A, Nurk S, Antipov D, Gurevich AA, Dvorkin M, Kulikov AS, et al. SPAdes: a new genome assembly algorithm and its applications to single-cell sequencing. J Comput Biol. 2012;19(5):455–77.

17. T.S. Prokka: rapid prokaryotic genome annotation. Bioinformatics. 2014;30:2068–9.

18. Gupta SK, Padmanabhan BR, Diene SM, Lopez-Rojas R, Kempf M, Landraud L, et al. ARG-ANNOT, a new bioinformatic tool to discover antibiotic resistance genes in bacterial genomes. Antimicrob Agents Chemother. 2014;58(1):212–20.

19. Carattoli A, Zankari E, Garcia-Fernandez A, Voldby Larsen M, Lund O, Villa L, et al. In silico detection and typing of plasmids using PlasmidFinder and plasmid multilocus sequence typing. Antimicrob Agents Chemother. 2014;58(7):3895–903.

20. Siguier P, Perochon J, Lestrade L, Mahillon J, Chandler M. ISfinder: the reference centre for bacterial insertion sequences. Nucleic Acids Res. 2006;1(34).

21. Chen L, Yang J, Yu J, Yao Z, Sun L, Shen Y, et al. VFDB: a reference database for bacterial virulence factors. Nucleic Acids Res. 2005;33(Database issue):D325–8.

22. Letunic I, Bork P. Interactive tree of life (iTOL) v3: an online tool for the display and annotation of phylogenetic and other trees. Nucleic Acids Res. 2016;44(W1):W242–5.

23. Sinha S, Sahu S, Pati J, Ray B, Pattnaik SK. Retrospective analysis of colistin-resistant bacteria in a tertiary care centre in India. Indian J Med Res. 2019;149(3):418–22.

24. Shankar C, Venkatesan M, Rajan R, Mani D, Lal B, Prakash JAJ, et al. Molecular characterization of colistin-resistant Klebsiella pneumoniae & its clonal relationship among Indian isolates. Indian J Med Res. 2019;149(2):199–207.

25. Aghapour Z, Gholizadeh P, Ganbarov K, Bialvaei AZ, Mahmood SS, Tanomand A, et al. Molecular mechanisms related to colistin resistance in Enterobacteriaceae. Infect Drug Resist. 2019;12:965–75.

26. Haeili M, Javani A, Moradi J, Jafari Z, Feizabadi MM, Babaei E. MgrB Alterations Mediate Colistin Resistance in Klebsiella pneumoniae Isolates from Iran. Frontiers in microbiology. 2017;8:2470.

27. Mathur P, Veeraraghavan B, Devanga Ragupathi NK, Inbanathan FY, Khurana S, Bhardwaj N, et al. First Report on a Cluster of Colistin-Resistant Klebsiella pneumoniae Strains Isolated from a Tertiary Care Center in India: Whole-Genome Shotgun Sequencing. Genome announcements. 2017;5(5).

28. Pragasam AK, Shankar C, Veeraraghavan B, Biswas I, Nabarro LE, Inbanathan FY, et al. Molecular Mechanisms of Colistin Resistance in Klebsiella pneumoniae Causing Bacteremia from India-A First Report. Frontiers in microbiology. 2016;7:2135.

29. Long SW, Olsen RJ, Eagar TN, Beres SB, Zhao P, Davis JJ, et al. Population Genomic Analysis of 1,777 Extended-Spectrum Beta-Lactamase-Producing Klebsiella pneumoniae Isolates, Houston, Texas: Unexpected Abundance of Clonal Group 307. mBio. 2017;8(3).

30. Lomonaco S, Crawford MA, Lascols C, Timme RE, Anderson K, Hodge DR, et al. Resistome of carbapenem-and colistin-resistant Klebsiella pneumoniae clinical isolates. PLoS One. 2018;8(13):e0198526.

31. Wyres KL, Nguyen TNT, Lam MMC, Judd LM, Chau NvV, Dance DAB, et al. Genomic surveillance for hypervirulence and multi-drug resistance in invasive Klebsiella pneumoniae from South and Southeast Asia. Genome Medicine. 2020;12(11).

32. Lam MMC, Wick RR, Wyres KL, Gorrie CL, Judd LM, Jenney AWJ, et al. Genetic diversity, mobilisation and spread of the yersiniabactin-encoding mobile element ICEKp in Klebsiella pneumoniae populations. Microb Genom. 2018;4(9):e000196.

33. Malhotra-Kumar S, Xavier BB, Das AJ, Lammens C, Butaye P, Goossens H. Colistin resistance gene mcr-1 harboured on a multidrug resistant plasmid. lancet Infect Dis. 2016;16(3):283–4.

34. Sekyere JO, Amoako DG. Genomic and phenotypic characterisation of fluoroquinolone resistance mechanisms in Enterobacteriaceae in Durban, South Africa. PLoS One. 2017;12(6):e0178888.

